# Utah-Stanford Ventilator (Vent4US): Developing a rapidly scalable ventilator for COVID-19 patients with ARDS

**DOI:** 10.1101/2020.04.18.20070367

**Authors:** Hongquan Li, Ethan Li, Deepak Krishnamurthy, Patrick Kolbay, Beca Chacin, Soeren Hoehne, Jim Cybulski, Lara Brewer, Tomasz Petelenz, Joseph Orr, Derek Sakata, Thomas Clardy, Kai Kuck, Manu Prakash

**Author notes:** co-first authors (Hongquan Li, Ethan Li, Deepak Krishnamurthy, Patrick Kolbay).

## Abstract

We describe a minimum, rapidly scalable ventilator designed for COVID-19 patients with ARDS. Our design philosophy is not only to try to address potential ventilator shortages, but also to account for uncertainties in the supply chains of parts commonly used in traditional ventilators. To do so we employ a modular design approach and broadly explore taking advantage of parts from non-traditional supply chains. In our current prototype, we demonstrate volume control with assist control on a test lung and present a linear actuator-driven pinch valve-based implementation for both pressure control and volume control with decelerating inspiratory flow. We estimate the component cost of the system to be around $500. We publish our draft design documents and current implementation which is open and accessible in the hope that broadening the community globally will accelerate arriving at a solution and that peer review will improve the final design.

## 1. Introduction

The current global pandemic of the COVID-19 respiratory disease caused by the SARS-CoV-2 virus has spread rapidly, with a total of 2,244,000 confirmed cases and more than 154,000 deaths as of 4/17/2020 [1]. Early reports suggest that up to 20% of people infected with SARS-CoV-2 develop severe disease requiring hospitalization, with up to 25% of hospitalized patients requiring intensive care unit (ICU) admission [2]. Many of the COVID-19 patients admitted to the ICU present with hypoxemic respiratory failure due to severe acute respiratory distress syndrome (ARDS), the clinical management of which involves intubation and mechanical ventilation which can last two weeks.

Both in the U.S. and globally, there is a shortage of ventilators for COVID-19 patients with severe ARDS. Present estimates of U.S. ventilator capacity include 62,000 fully-featured modern ventilators, 98,000 ventilators with more limited capabilities, and only 12,700 ventilators in national stockpiles for emergency purposes [3, 4]. Not only is the number of critical care ventilators limited, even the production capacity of the global medical device industry is insufficient to meet demand [5]. Furthermore, many low-capacity healthcare settings across the world are projected to face extreme shortages of ventilators and ICU beds for COVID-19 patients: recent reports found that the Central African Republic (pop. 4.8M) has a total of three suitable ventilators [6], while South Sudan (pop. 11.7M) has four ventilators [7], Northeast Syria has 11 ventilators [7], Northwest Syria (pop. 3M) has 30 adult ventilators [7], Burkina Faso (pop. 20.9M) and Sierra Leone (pop. 7.9M) both have between 10 and 15 ventilators [7], Indonesia (pop. 267M) has approximately 8400 ventilators [8], and India (pop. 1.3B) has approximately 20,000 ventilators [9].

To address the global shortage of ventilators, we explore the possibility of developing a ventilator which is minimal yet designed for ventilating COVID-19 patients with ARDS. In doing so, we also hope to provide a modular and accessible reference design which may help accelerate efforts in finding solutions. We use this document to share our designs and findings.

## 2. Clinical requirements and their design implications

From examining guidelines published by various governmental agencies [10–12] and non-governmental organizations [13–17], reviewing clinical literature [2, 18–25], talking to clinicians, and reviewing system requirements and specifications from other ventilator projects [26–28], we have identified the following overall minimum requirements to guide our design:

1. the ability to achieve lung-protective ventilation with low tidal volume (4–8 mL/kg predicted body weight)
2. the ability to measure and keep inspiratory plateau pressure not exceeding 30 cmH2O, as part of the lung protective strategy
3. the ability to set positive end-expiratory pressure (PEEP), to maximize and maintain alveolar recruitment for improved oxygenation
4. the ability to provide mechanical support for patient-initiated breaths
5. the ability to use respiratory rates of up to 30 breaths/minute
6. sufficient safety measures to lower the risk of barotrauma, rebreathing, hypoxia, and hypoventilation, including alarms to draw attention of clinical personnel to untoward situations including disconnect or low pressure

Requirements (1)-(4) and (6) imply that flow and airway pressure need to be measured. Requirement (5) implies support of high inspiratory flow rate and fast exhalation. In addition to the above minimum requirements, the following capabilities will enhance the ventilator:

1. both volume control mode and pressure control mode, to allow clinicians to choose a mode based on their training and preference
2. decelerating inspiratory waveform in volume control mode, to distribute the inspiratory tidal volume more evenly in heterogeneous lung conditions
3. pressure support ventilation mode

These requirements imply that a proportional valve capable of regulating flow needs to be used for controlling inspiration.

Other requirements include the ability to set the fraction of inspired oxygen (FiO2) and confirm FiO2 through measurement. System integration of FiO2-related functionality will be demonstrated in a future update.

For the full list of identified requirements for a minimally viable ventilation system for COVID-19 patients, including requirements on operating ranges of settings and parameters, display and accuracy of measurements, safety protections and alarms to mitigate potential hazards, reliability, and ease of use, please refer to Supplementary Text 1.

## 3. Results

Through rapid iterative development, we arrived at our current tentative design shown in Supplementary Figures S1 and S2, with concrete realization as a prototype design shown in Figure 1.

**Fig. 1.**
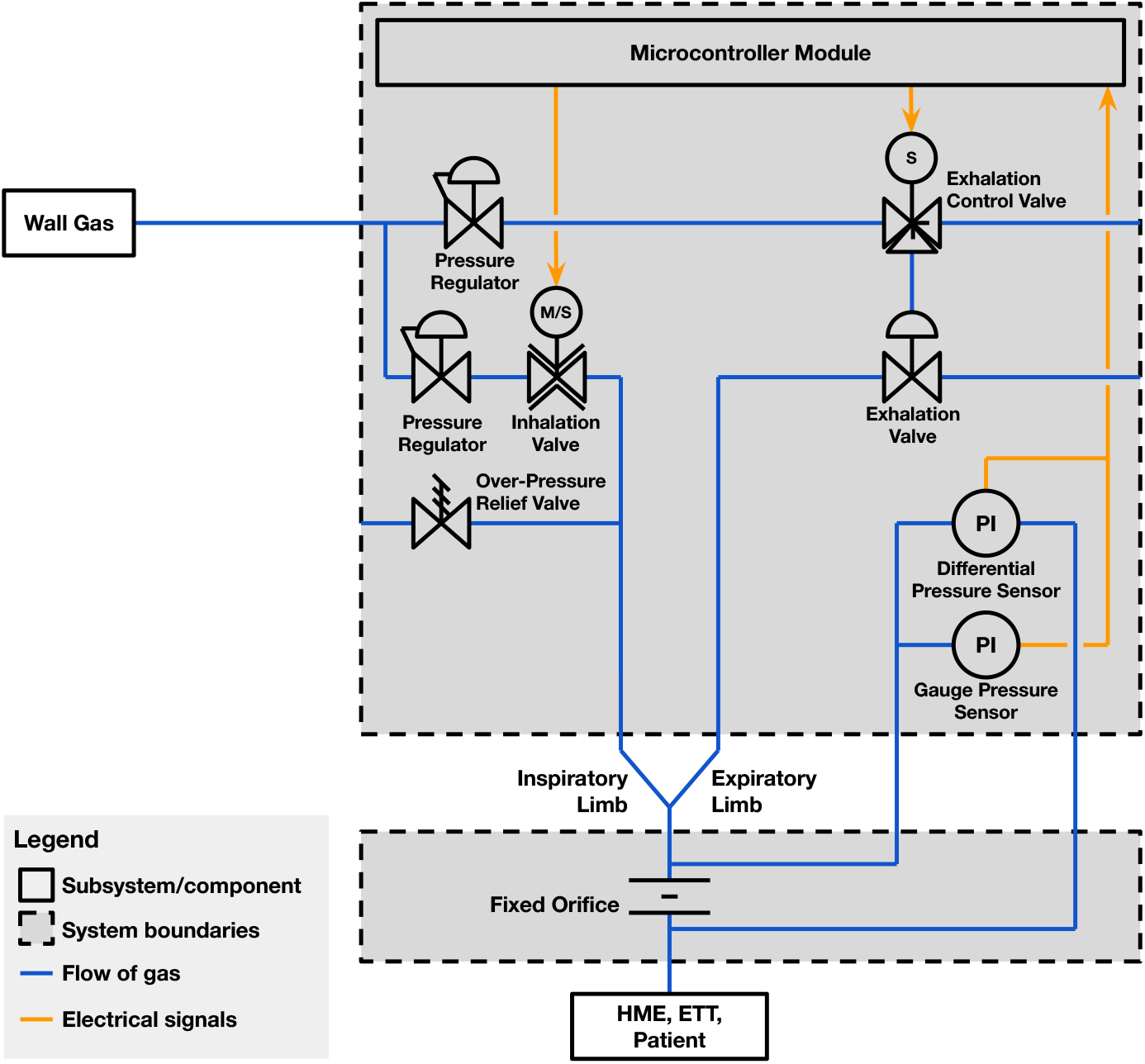
Simplified piping and instrumentation diagram (P&ID) schematic of the current breathing circuit prototype. The full proposed design is shown in Supplementary Figures S1 and S2.

The system uses compressed medical air and oxygen which can be provided from either central hospital supply lines or cylinders. For the simplest achievable design, we present square wave volume control with the inspiratory valve consisting of a solenoid pinch valve found in microfluidics/analytical instruments. For expiration, we use a three-way solenoid pinch valve to pneumatically control an exhalation valve already used in existing breathing circuits. A differential pressure sensor is used with an off-the-shelf flow transducer (Hamilton Medical AG, Bonaduz, Switzerland) placed close to the breathing circuit’s Y-piece to measure flow and calculate inhaled and exhaled volume, and a vented gauge pressure sensor is used to monitor the airway pressure. PEEP can be controlled electronically by closing the exhalation valve when the airway pressure drops below the set value. A microcontroller is used to achieve real-time control of the system, with an internal timer driving sensor readings at a 2.5 ms interval. For displaying waveforms and setting ventilator parameters in the prototype system, the microcontroller talks to a computer using a USB serial interface through a virtual COM port at 2 Mbps. The computer runs a PyQt-based program with a graphical user interface which we have implemented in Python. The code for both the microcontroller firmware and the graphical user interface software can be found at https://github.com/vent4us/vent-dev. For deployment, possible human-machine interface options include but are not limited to a Wio Terminal (Seeed Studio, Shenzhen, China), a Raspberry Pi with touchscreen (Raspberry Pi Foundation, Cambridge, United Kingdom), and an NVIDIA Jetson Nano (NVIDIA, Santa Clara, CA) with touchscreen. So far we have functionally tested the implementation based on the Jetson Nano. Photos of the prototyping system and core components used in the prototype are shown in Figure 2. The total cost of the core components for this minimum system is around $200 (excluding the battery).

**Fig. 2.**
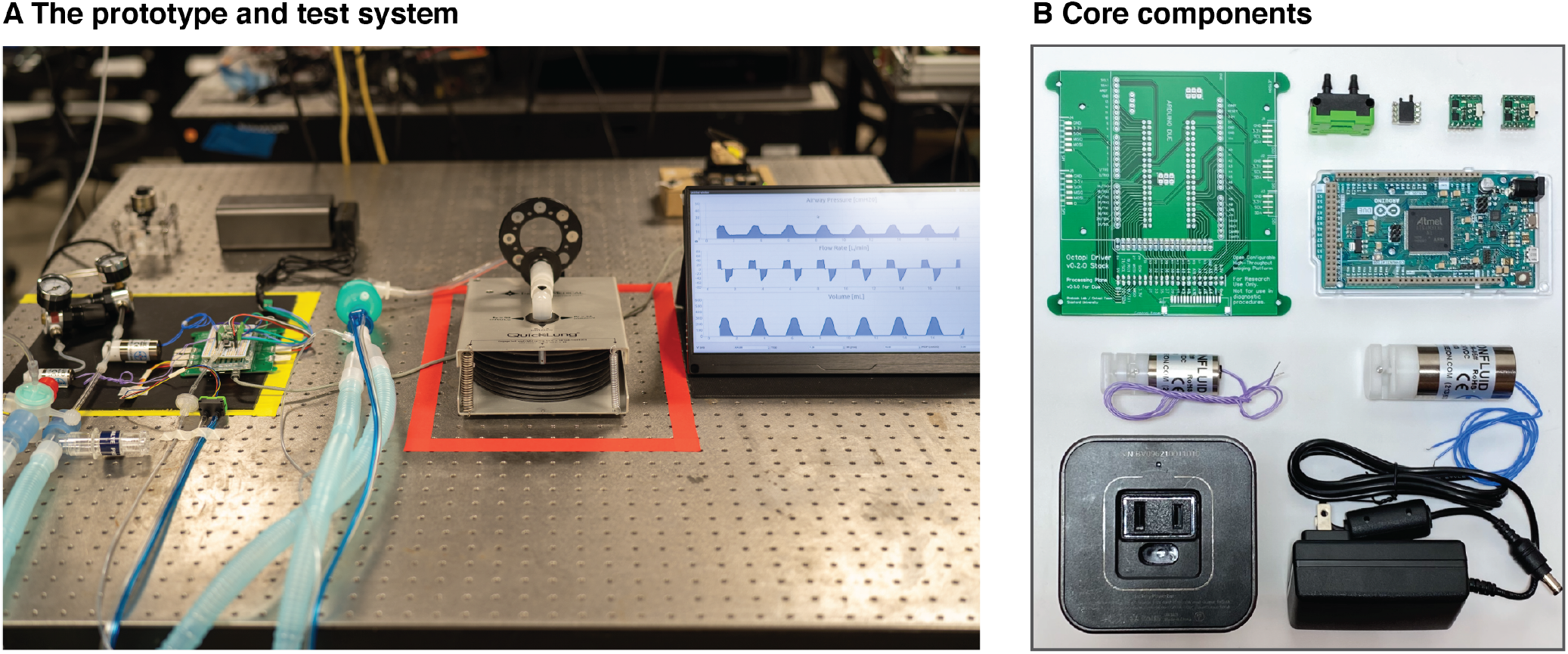
The prototype and test system are shown in (A), with the ventilator components in the area outlined by yellow tape, and the test lung in the area outlined by red tape. The core components used in the prototype system, shown in (B), include a differential pressure sensor, a vented gauge pressure sensor, two MOSFETs for controlling solenoids, an Arduino Due microcontroller board, a three-way solenoid pinch valve (1/8” OD), and a two-way solenoid pinch valve (1/4” OD). In the test system, an exhalation valve from the IPPB breathing circuit and a Hamilton Medical flow transducer are used.

As part of the development process and for initial tests, we connected the system to a test lung (QuickLung, Ingmar Medical, Pittsburgh, PA). Figure 3 shows waveforms demonstrating control of tidal volume, PEEP, respiratory rate, as well as assist control where a breath of set tidal volume can be triggered by the patient.

**Fig. 3.**
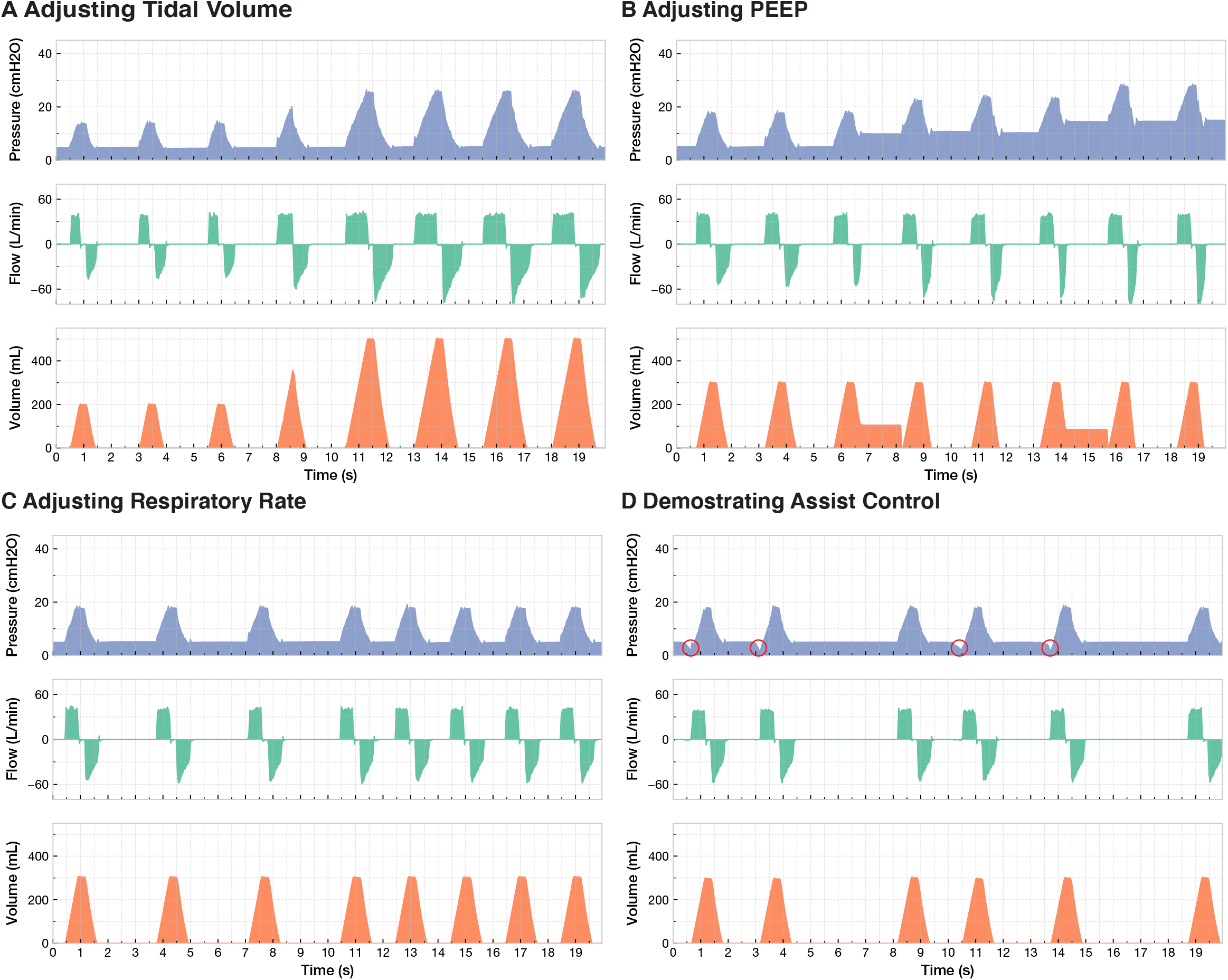
Waveforms showing (A) adjustments of tidal volume (from 200 mL to 500 mL), (B) adjustments of PEEP (5 cmH2O, 10 cmH2O and 15 cmH2O), (C) adjustments of respiratory rate (18 breaths/min to 30 breaths/min), and (D) patient-triggered breaths in assist control (pressure drops are artificially generated to simulate the intubated patient trying to initiate breath). In these waveforms, the displayed pressure includes compensation for resistance due to the flow transducer, the HME filter and the endotracheal tube. Example waveforms without this compensation are shown in Supplementary Figure S3.

The linear actuator (Figure 4A) compresses a segment of 1/4” OD, 1/8” ID silicone tubing (hardness: Shore A45) by variable amounts to achieve flow control. At an inlet pressure of 15 psi, we found that a 3 mm actuation is sufficient for valve flow to transition from a completely closed state to a completely open state (Figure 4B and 4C), with the completely open state allowing the maximum flow rate of 50 L/min. We also demonstrated both slow and fast actuation cycles of the valve (Figure 4D and 4E, at actuation rates for fully-closed to fully-open cycles up to 5 Hz). We measured the valve’s response time (time it takes for flow to reach 95% of its maximum value) to be 48 ± 8 ms (Figure 4F). Finally we tested the repeatability of the valve’s proportional flow control by measuring the flow rate as a function of valve state (Figure 4G). The valve was found to be fairly repeatable, with the flow rate at a given valve state having a maximum variability of around 5% at the highest flow rates. Repeatability can be further improved by incorporating an encoder or a Hall effect limit switch.

**Fig. 4.**
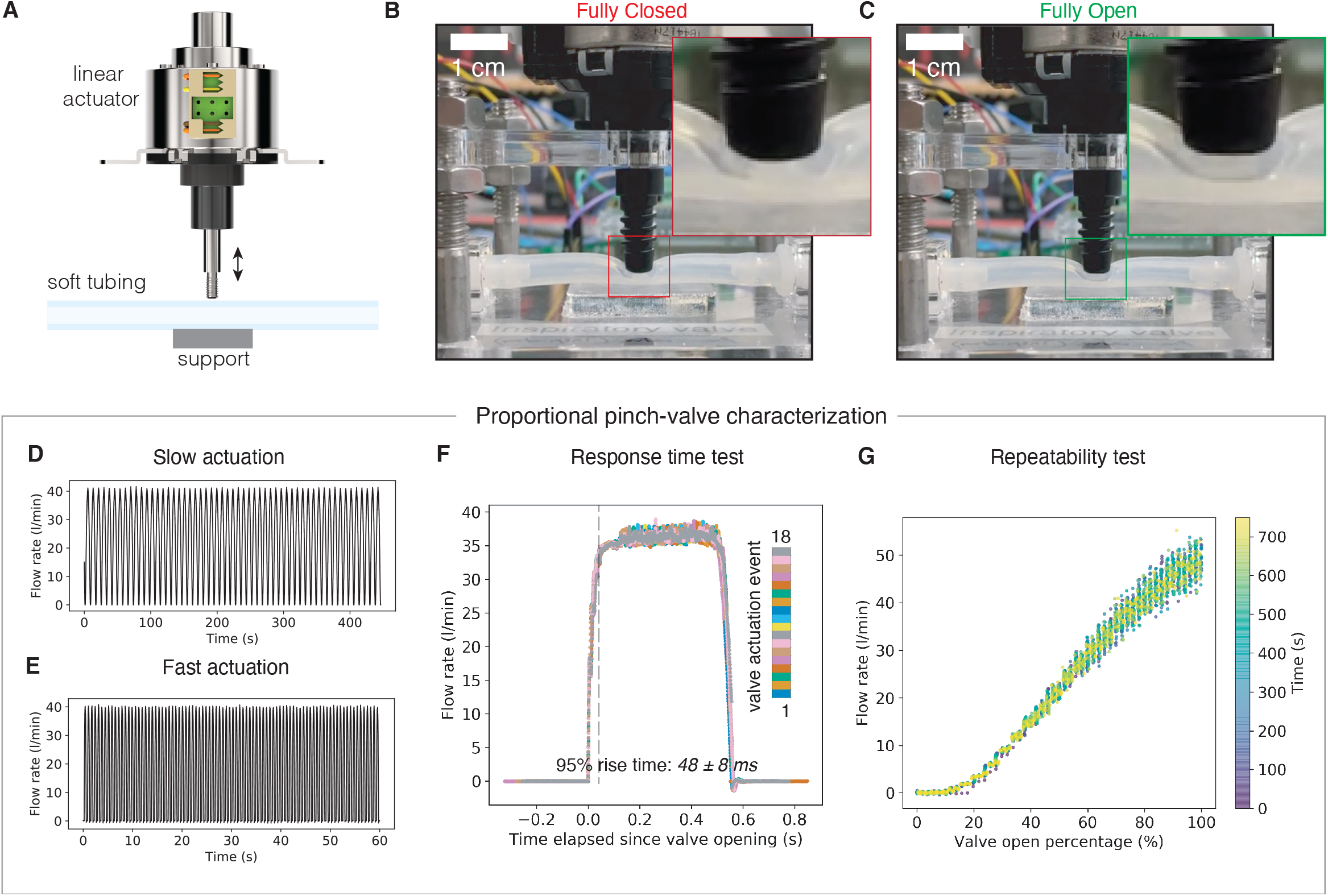
Design, prototyping, and characterization of a linear actuator-driven pinch valve. (A) Schematic of the pinch valve design using a ubiquitous automotive headlamp linear actuator. (B), (C) Photographs of the valve prototype, in the fully-open and fully-closed states, respectively. (D), (E) Demonstration of slow and fast valve actuation cycles. (F) Rise-time dynamics of flow rate when valve is used as an ON-OFF valve actuated from fully-closed to fully-open flow state. (G) Same valve used in proportional control mode showing the repeatability of flow rate vs valve position.

As a proof-of-concept demonstration, we replaced the solenoid pinch valve with our homemade linear actuator-driven pinch valve (Figure 5) and implemented decelerating inspiratory flow. Two resulting waveforms are shown in Figure 6.

**Fig. 5.**
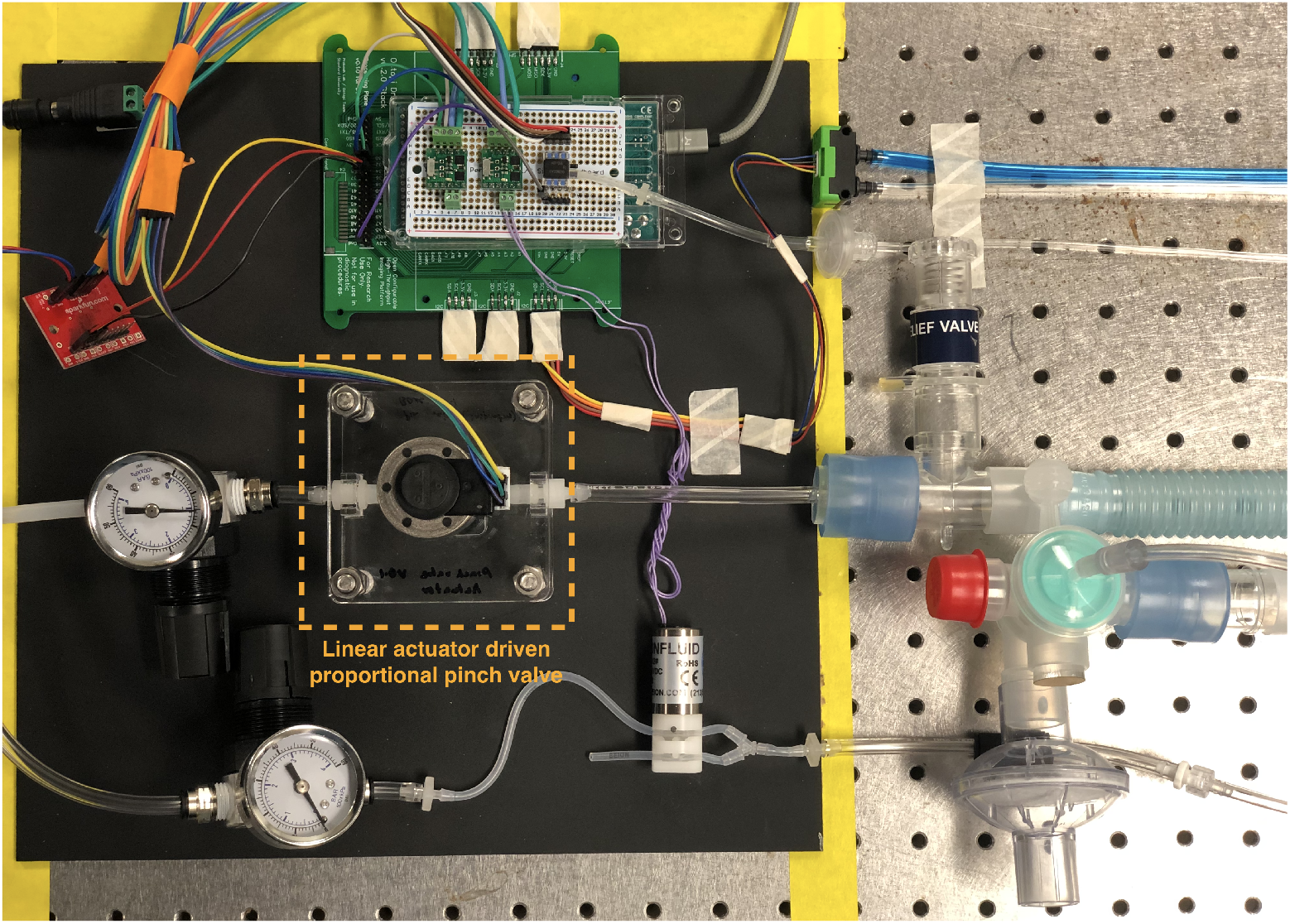
Linear actuator-driven pinch valve in the prototype, in place of the two-way solenoid pinch valve as the inspiratory valve.

**Fig. 6.**
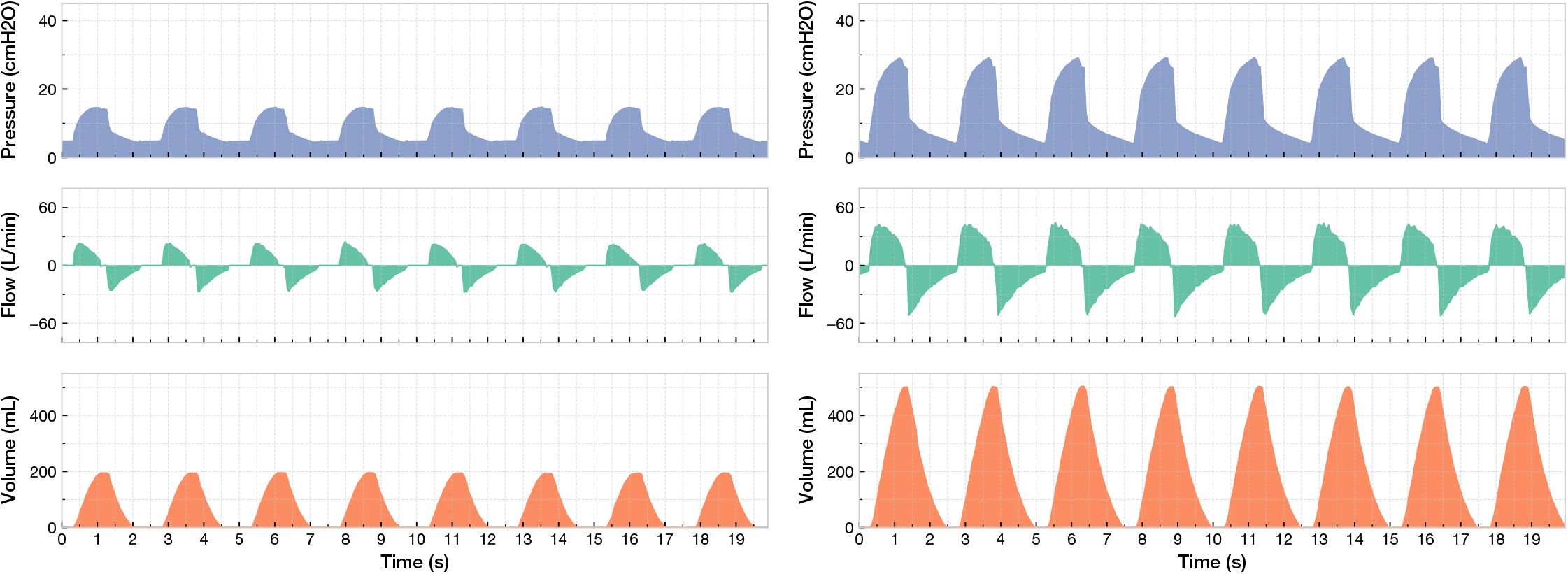
Waveforms from the configuration where the linear actuator-driven pinch valve is used as the inspiratory valve to achieve decelerating inspiratory flow. PEEP is 5 cmH2O, tidal volume is 200 mL (left) and 500 mL (right).

## 4. Discussion

We have developed a prototype of a minimum ventilator specifically designed to provide the capabilities needed for ventilating COVID-19 patients with ARDS. Next steps include implementing and testing pressure control and pressure support with linear actuator-driven proportional pinch valves, more in-depth characterization of the system in terms of the parameter space that it can operate in, and completing the system with other components which remain to be integrated (Supplementary Figure S2). Subsequently, we will finish design for safe and reliable long-term operation of the system in order to meet the associated requirements for hardware and software.

We will be releasing all our designs in the open-source domain very soon and will be uploading content files with an open-source license shortly at http://www.vent4us.org.

## Data Availability

All data is included in the manuscript. All code developed for this work is openly accessible at out GitHub repository at https://github.com/vent4us/vent-dev

http://www.vent4us.org

## Acknowledgements

We thank Brian Daniel at UCSF for consultation and feedback. We thank all members of Prakash Lab and of the University of Utah Anesthesiology Bioengineering Lab for feedbacks. We thank Schmidt Futures, Moore Foundation, CZI BioHub and HHMI-Gates Foundation for financial support to the Prakash Lab.

## Supplementary Text 1: System Requirements

Our current understanding is that a minimally viable ventilation system must provide the following required (**must-have**) functionalities for target users:

1. Continuous Mandatory Ventilation (CMV) with pressure control and volume monitoring for one intubated patient:
  a. Interface to ISO 5359:2014 and/or ISO 18082:2014-standard connectors to externally-provided medical air and O2 sources supplied through wall flow limiters outputting at a total flow rate between 20 - 25 L/min and pressures of up to 65 psi.
  b. Interface to ISO 5367:2014 and ISO 5356-1:2015-standard connectors for invasive ventilation.
  c. Compatibility with any FiO2 level between 21% - 100% as adjusted by flow ratios through wall rotameter settings.
  d. Manual adjustment of ventilation settings to indirectly achieve tidal volumes across the entire range of 250 - 800 mL for patients with any lung compliance over 30 mL/cmH2O while maintaining plateau pressure ≤30 cmH2O.
  e. Airway pressurization to at least the PEEP level setting at all times.
  f. Feasibility of inspiration at peak flow rate of 120 L/min and expiration at peak flow rate of 85 L/min.
  g. Ventilation support for patient-initiated breaths.
2. Adjustment of the settings listed in Table 1 across the required ranges, with minimum accuracies (upper bounds on allowed differences between user inputs and system behavior), maximum granularities (upper bounds on smallest provided increments of adjustment), default values, and display requirements:
3. Bedside monitoring with continuous display of achieved measurements of the quantities listed in Table 2, with minimum accuracies over the ranges listed and indications when values are beyond sensor ranges:
4. Safety protections to minimize the likelihood of entering the following hazardous failure states:
  a. Excessive airway pressure, with the threshold for “excessive pressure” at 45 cmH2O, with an accuracy of ± 5 cmH2O.
  b. Failure of software.
  c. Emission of RF or EM signals which could interfere with other critical care equipment.
  d. Significant deterioration of ventilation performance due to RF or EM emission.
  e. Disruption of ventilation functionality within 30 minutes after loss of external power.
  f. Disruption of alarm functionality within 60 minutes after loss of external power.
  g. Accumulation of sensor drift leading to loss of required measurement accuracies.
  h. Partial or full blockage of pressure lines leading to loss of required ventilation performance.
  i. Patient rebreathing of exhaled CO2 at an inspiratory concentration >1%
  j. Apnea when the patient attempts to initiate a breath, with the threshold for “apnea” at an airway inspiratory flow rate of ≤60 L/min while airway pressure < −2 cmH2O.
  k. Contamination of the patient’s airway.
  l. Fire.
5. In-room and out-of-room alarms, noticeable from a distance of at least 10 feet away, upon entering the following exceptional error states:
  a. Ventilation:
    i. Measured pressure of medical air or O2 supply outside range allowed by 1.a.
    ii. Measured airway pressure at 0 cmH2O during ventilation, for example resulting from disconnection of gas lines.
    iii. Measured airway pressure outside user-adjustable lower and upper limits.
    iv. Measured PIP outside user-adjustable lower and upper limits.
    v. Measured plateau pressure outside user-adjustable lower and upper limits.
    vi. Measured airway pressure during expiration above user-adjustable upper limit.
    vii. Measured PEEP outside user-adjustable lower and upper limits.
    viii. Measured tidal volume outside user-adjustable lower and upper limits.
    ix. Apnea over a user-adjustable minimum threshold, for example resulting from any failure to continue normal ventilation.
    x. Commanded system power-off during ventilation.
  b. Power:
    i. Loss of external power supply for longer than 15 s, such as would be caused by disconnection or failure of wall power combined with subsequent failure to restore wall power or switch to a portable external power source.
    ii. Depletion of internal backup power reserves for ventilation below 50% of full capacity.
    iii. Depletion of internal backup power reserves for alarms below 50% of full capacity.
  c. Other detectable failures of software or hardware subsystems or components.
6. Biological safety of all components in contact with breathing gases.
7. Infection control to prevent nosocomial infections:
  a. Disposability or decontaminability of all parts which come into contact with the patient’s breath.
  b. Impermeable casing around all working components of the device.
  c. Easy cleaning of all external surfaces after exposure to respiratory secretions or blood splatter.
  d. Pressure delivered must account for any pressure drop from an HME viral filter placed in-line on the endotracheal tube between the system and the patient.
  e. Controllable “stand-by” or “switch-off” functionality to stop flow during disconnection of ETT without aerosolization.
8. Reliability:
  a. Continuous ventilation at 100% duty cycle for 14 days without any subsystem or component failures and without servicing or replacement of any subsystems or components.
  b. Continuous operation without subsystem/component failures resulting from mechanical shocks and vibrations as encountered during regular patient transfer within a medical facility.
  c. Fast and easy servicing and replacement of subsystems/components to minimize downtime.
9. Ease of use:
  a. Portable, including single-handed carrying.
  b. Operation from standard 120 V or 240 V wall outlets for electrical power.
  c. Clear and intuitive user interface for adjustment of ventilation settings, monitoring of measured quantities, and identification of exceptional error states.
  d. Monitoring of internal power reserves for ventilation and alarm functionalities.
  e. Recharging of internal power reserves while external power is available.
  f. Continuous ventilation with simultaneous recharging of internal power reserves for 180 min from a portable external power source consisting of a fully-charged battery with a standard AC outlet.
  g. Delivery of ventilation while placed on a patient or patient bed.
  h. Easy adjustment of parameters by a user wearing full protective gear.
  i. Integrated clearly-marked usage instructions and labels of critical functions and controls.
  j. Easy access to parts for servicing or replacement by a technician. Based on our current understanding, the ventilation system may or may not provide any of the following optional (**nice-to-have**) functionalities for target users, depending on whether these can be achieved with rapid development and large-scale rapid manufacturing:
10. Ventilation functionality:
  a. CMV with support for patient-initiated breaths.
  b. CMV with volume control.
  c. Pressure Support Ventilation (PSV) mode with failsafe to CMV if the patient stops breathing in PSV mode and real-time confirmation of each patient’s breath.
  d. Compatibility with the use of active humidification and viral filters as a substitute for the heat and moisture exchanger (HME) in the breathing circuit.
  e. Ventilation and ventilation control from an oxygen cylinder in settings where compressed medical air is not available.
11. Adjustment of the following parameters:
  a. Ventilatory frequency with fixed settings in increments of up to 2 bpm rather than up to 5 bpm
  b. On-board adjustment of FiO2, with FiO2 setting at a maximum allowed granularity of 10% over the entire range from 21% - 100%.
12. Bedside monitoring with continuous display of achieved measurements of the following quantities:
  a. Pressures of the medical air and O2 sources provided to the system.
  b. Waveforms of actual airway pressure, measured flow, and measured volume.
13. Reliability:
  a. Diagnostic logging of faults and failures in the system for post-marketing surveillance and tracking of safety margins from successful handling of faults in subsystems or components.
  b. Reliable operation for at least 10 patients.
14. Usability:
  a. “Far-View” capability, i.e., one or two critical device parameters are visible from up to 5 yards away (to allow ascertaining basic ventilator function from outside of the room without having to don/doff PPE).
  b. USB communication interface to an external device such as a laptop computer.
  c. Remote adjustment of the parameters described by items 2, 13, and 14.
  d. Remote monitoring of the quantities described by items 3, 14, 16, and 17.

**Table S1:** Requirements for adjustable settings for ventilation.

| Req. ID | Setting | Range | Accuracy | Granularity | Default Value | Continuous Display |
| --- | --- | --- | --- | --- | --- | --- |
| 2.a | Respiratory Rate | 10 - 30 bpm | $\pm 2$ bpm | 2 bpm | None | Required |
| 2.b | I:E Ratio | 1:3 - 3:1 | $\pm 0.1$ s | 0.2 I, 0.2 E | 1:2 | Optional |
| 2.c | PIP | 5 - 40 cmH<sub>2</sub>O | $\pm 2$ cmH<sub>2</sub>O | 5 cmH<sub>2</sub>O | None | Required |
| 2.d | PEEP | 5 - 20 cmH<sub>2</sub>O | $\pm 2$ cmH<sub>2</sub>O | 5 cmH<sub>2</sub>O | None | Required |
| 2.e | Inspiratory Time | 0.5 - 4.5 s | $\pm 0.1$ s | 0.2 s | 0.4-0.5 | Optional |

**Table S2:** Requirements for continuous display of real-time achieved measurements of ventilation.

| Req. ID | Measurement | Range for Required Accuracy | Accuracy |
| --- | --- | --- | --- |
| 3.a | Respiratory Rate | 10 - 30 bpm | $\pm 2$ bpm |
| 3.b | Tidal Volume | 350 - 1000 mL | 10% |
| 3.c | PIP | 5 - 40 cmH<sub>2</sub>O | $\pm 2$ cmH<sub>2</sub>O |
| 3.d | PEEP | 0 - 20 cmH<sub>2</sub>O | $\pm 2$ cmH<sub>2</sub>O |
| 3.e | FiO<sub>2</sub> | 21% - 100% | $\pm 5\%$ FiO<sub>2</sub> |
| 3.f | Airway Pressure | 0 - 40 cmH<sub>2</sub>O | $\pm 2$ cmH<sub>2</sub>O |

**Supplementary Fig. S1.**
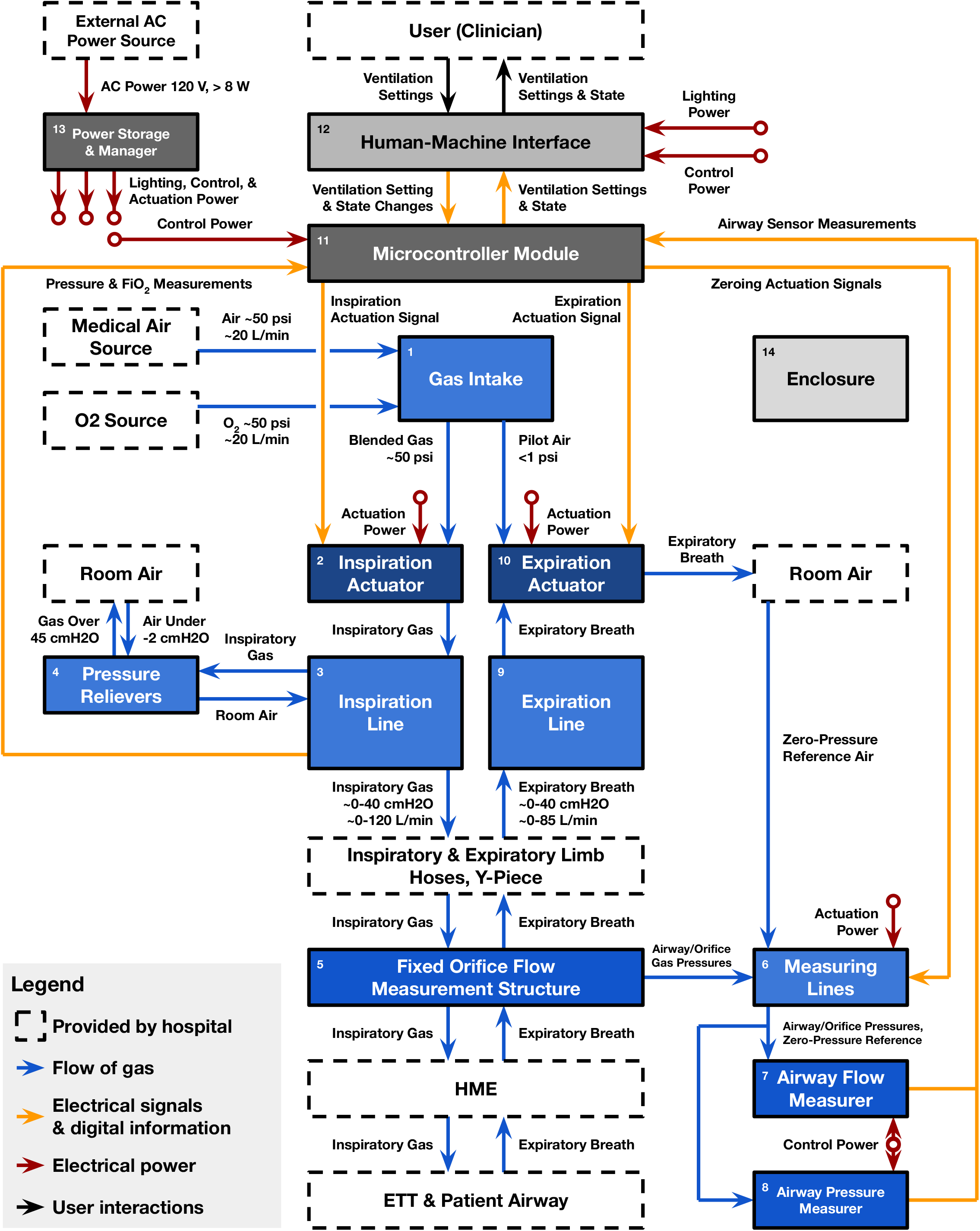
Functional block diagram for tentative overall system design. Solid blocks are subsystems to be provided by the system and will be described by design specifications. Dashed blocks are components provided by the hospital, and supported operating conditions involving them are described by the system requirements listed in Supplementary Text 1. Interactions between subsystems are represented by arrows labeled with order-of-magnitude estimates where available.

**Supplementary Fig. S2.**
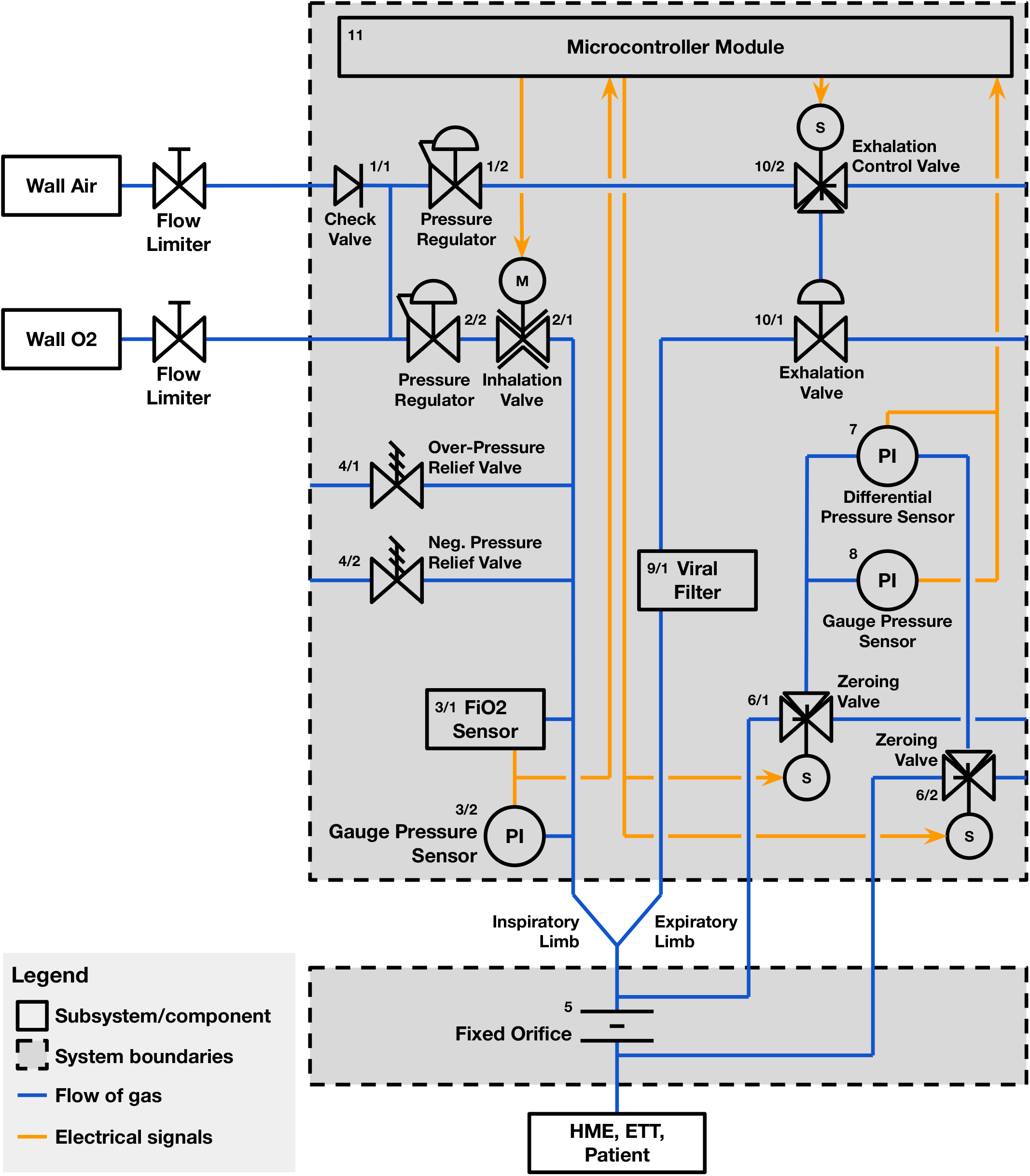
P&ID schematic of tentative full design of the breathing circuit to implement the overall system design shown in Supplementary Figure S1. Components shown in this diagram which are omitted from Figure 1 have not yet been characterized or integrated into the prototype breathing circuit.

**Supplementary Fig. S3.**
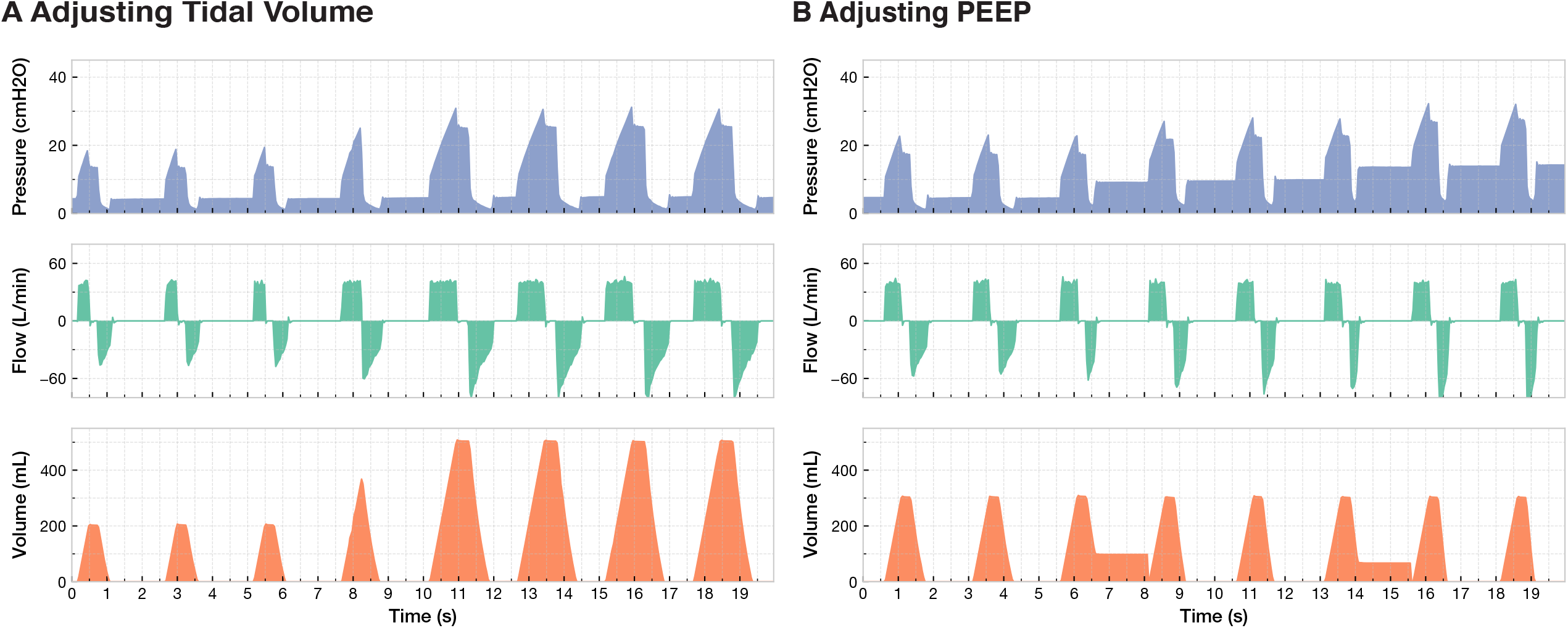
Waveforms obtained without compensating for pressure drop caused by resistance in the flow transducer, the HME filter, and the endotracheal tube. Note the presence of large spikes and dips visible in the waveforms when flow switches between roughly 40 L/min and 0 L/min, in contrast to the waveforms in Figure 3A and 3B.

## Notes

### Competing Interest Statement

The authors have declared no competing interest.

